# Pre-diagnostic circulating concentrations of insulin-like growth factor-1 and risk of COVID-19 mortality: results from UK Biobank

**DOI:** 10.1101/2020.07.09.20149369

**Authors:** Xikang Fan, Cheng Yin, Jiayu Wang, Mingjia Yang, Hongxia Ma, Guangfu Jin, Mingyang Song, Zhibin Hu, Hongbing Shen, Dong Hang

## Abstract

**Background:** Coronavirus disease 2019 (COVID-19) deteriorates suddenly primarily due to excessive inflammatory injury, and insulin-like growth factor-1 (IGF-1) is implicated in endocrine control of the immune system. However, the effect of IGF-1 levels on COVID-19 prognosis remains unknown.

**Objective:** To investigate the association between circulating IGF-1 concentrations and mortality risk among COVID-19 patients.

**Design:** Prospective analysis.

**Setting:** UK Biobank.

**Participants:** 1425 COVID-19 patients who had pre-diagnostic serum IGF-1 measurements at baseline (2006-2010).

**Main outcome measures:** COVID-19 mortality (available death data updated to 22 May 2020). Unconditional logistic regression was performed to estimate the odds ratio (OR) and 95% confidence intervals (CIs) of mortality across the IGF-1 quartiles.

**Results:** Among 1425 COVID-19 patients, 365 deaths occurred due to COVID-19. Compared to the lowest quartile of IGF-1 concentrations, the highest quartile was associated with a 37% lower risk of mortality (OR: 0.63, 95% CI: 0.43-0.93, *P*-trend=0.03). The association was stronger in women and nonsmokers (both *P*-interaction=0.01).

**Conclusions:** Higher IGF-1 concentrations are associated with a lower risk of COVID-19 mortality. Further studies are required to determine whether and how targeting IGF-1 pathway might improve COVID-19 prognosis.

## Introduction

An outbreak of Coronavirus Disease 2019 (COVID-19) began in December 2019 and has triggered a public health emergency of international concern ^1^. The majority of COVID-19 patients have a self-limiting infection and recover, but some suffer severe symptoms and even die ^2^. The suddenly deteriorating conditions of some patients are mainly attributed to systematic inflammatory injury caused by the excessive or uncontrolled activation of immune response, known as the cytokine storm, leading to respiratory distress syndrome (ARDS) and multiple organ failure ^3^.

Insulin-like growth factor 1 (IGF-1) belongs to the IGF family that plays a critical role in diverse biological activities including cell proliferation, metabolism, differentiation, and survival ^4^. Cumulative evidence also supports that IGF-1 pathway can regulate the immune response via interaction with various cytokines (e.g., interferons) and immune cells such as T lymphocytes, macrophages, and bone marrow cells ^5^. IGF-1 may act as an important switch controlling the amplitude and quality of the immune response ^6^. Of note, IGF-1 administration is already an approved therapy for growth failure and may improve symptoms of autoimmune diseases such as type-1 diabetes and multiple sclerosis ^7^. However, whether IGF-1 plays a role in COVID-19 prognosis remains unknown.

Therefore, in the current study, we used the UK Biobank resource, with recently released data on COVID-19 tests and updated death records, to investigate the association between pre-diagnostic serum IGF-1 concentrations and mortality among COVID-19 patients.

## Methods

### Study Participants

UK Biobank is a prospective cohort study consisting of approximately half a million participants (aged 37-73 years) recruited across the UK between 2006 and 2010 ^8^. Up to 22 May 2020, among 5516 participants who had undergone COVID-19 PCR tests provided by Public Health England, 1422 patients were diagnosed with COVID-19. In the current analysis, we also included additional 150 patients who died from COVID-19 according to the National Health Service death records. The patients who had no available data on serumIGF-1 (n=140) or died from other causes (n=7, including colon cancer, lung cancer, dementia, chronic ischaemic heart disease, and bronchopneumonia) were excluded, leaving 1425 patients in the final analysis.

### Assessment of IGF-1

Details about serum biomarker measurements and assay performances have been described online (http://biobank.ndph.ox.ac.uk/showcase/showcase/docs/serum_biochemistry.pdf). Briefly, serum concentrations of IGF-1 were measured in UK Biobank’s purpose-built facility using a ‘one step sandwich’ Chemiluminescent Immunoassay method based on DiaSorin Liaison XL Analyzer (Diasorin S.p.A), with a detection range of 1.3-195 nmol/L. The average coefficients of variation of IGF-1 derived from internal quality control samples of known high, medium, and low concentrations were 6.18%, 5.29%, and 6.03%, respectively.

Moreover, the assay of serum IGF-1 was registered with an external quality assurance (EQA) scheme (RIQAS Immunoassay Speciality 1) to verify accuracy. The EQA results showed that 100% of participated distributions (n=105) were good or acceptable.

### Ascertainment of COVID-19 Mortality

Dates and causes of death were obtained from death certificates held by the National Health Service Information Centre (England and Wales) and the National Health Service Central Register Scotland (Scotland) ^9^. The outcome of current study was mortality due to COVID-19 (ICD-10 U07) and available death data were updated to 22 May 2020.

### Ascertainment of Covariates

At baseline, participants attended one of 22 assessment centers across England, Scotland, and Wales where they completed a touch-screen, self-completed questionnaire. Ethnicity, smoking status, and alcohol intake were self-reported. Height and body weight were measured by trained nurses at baseline, and body mass index (BMI) was calculated as weight in kilograms divided by height in meters squared. Townsend deprivation index was derived from residence using data on car and home ownership, unemployment, and household overcrowding. Physical activity was measured as total metabolic equivalent task-minutes per week for all activity including walking, moderate and vigorous activity.

### Statistical Analysis

Because the date of COVID-19 testing did not represent the time of infection, the survival time for each patient could not be accurately estimated, leading to Cox regression models inapplicable in the current analysis. Therefore, we analyzed the association between IGF-1 and COVID-19 mortality using unconditional logistic regression models. Odds ratios (OR) and 95% confidence intervals (CI) for each quartile of IGF-1 were calculated, with the lowest quartile as the reference. Model 1 was adjusted for major covariates including age at infection, sex, and ethnicity; Model 2 was further adjusted for Townsend deprivation index, BMI, smoking status, alcohol drinking, and physical activity. We also additionally adjusted for C-reactive protein in those who had the inflammatory biomarker data (n=1418), and the results were essentially unchanged (data not shown).

Stratified analyses were conducted according to the median age at infection (<70, ≥70 years), sex (male, female), BMI (<30, ≥30 kg/m^2^), physical activity (≤median, >median), and smoking status (never, ever) in the fully-adjusted model. To investigate potential effect modification by these stratification variables, we used a likelihood ratio test comparing the models with and without interaction terms between IGF-1 concentrations and each of the stratification variables.

Sensitivity analyses were performed by excluding participants with baseline cancer or cardiovascular disease. We used SAS 9.4 for all analyses. All statistical tests were two-sided, and *P* <0.05 was defined as statistically significant.

## Results

Among 1425 COVID-19 patients, 365 deaths occurred due to COVID-19. The median time from baseline blood draw to COVID-19 testing was 11.1 years (interquartile range: 10.5-11.8 years). The distribution of IGF-1 concentrations is shown in **Figure 1**, ranging from 4.79 to 46.09 nmol/L, which was similar to the distribution in the whole cohort (data not shown).

**Figure 1.**
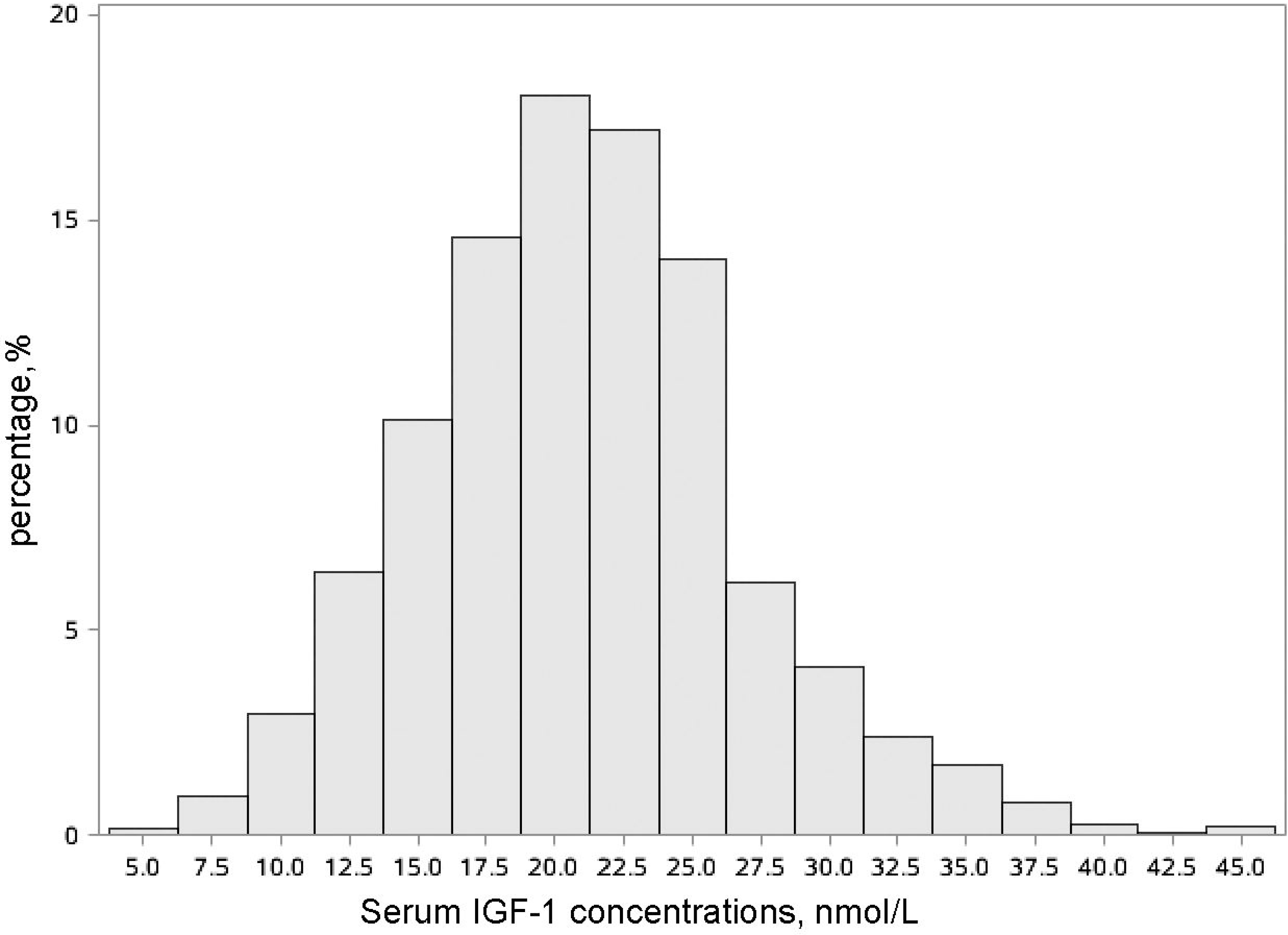
The distribution histogram of IGF-1 concentrations.

**Table 1** summarizes the main characteristics of COVID-19 patients according to quartiles of serum IGF-1 concentrations. At baseline, participants with higher IGF-1 had a lower BMI and higher levels of physical activity, and tended to be younger, males, and nonsmokers.

**Table 1.** Baseline characteristics of COVID-19 patients according to quartiles of serum IGF-1 concentrations.

| Characteristic | Serum IGF-1 concentrations, nmol/L |  |  |  |
| --- | --- | --- | --- | --- |
|  | Q1 (4.79-17.09) | Q2 (17.11-20.78) | Q3 (20.79-24.47) | Q4 (24.47-46.09) |
| Participants N | 356 | 356 | 357 | 356 |
| Age at blood draw, mean (SD), y | 60.08 (7.33) | 57.83 (9.24) | 56.52 (9.29) | 54.51 (9.36) |
| Age at infection, mean (SD), y | 71.26 (7.39) | 69.02 (9.36) | 67.64 (9.32) | 65.65 (9.32) |
| Female, No. (%) | 180 (50.56) | 168 (47.19) | 168 (47.06) | 150 (42.13) |
| White race, No. (%) | 315 (88.48) | 314 (88.20) | 299 (83.75) | 312 (87.64) |
| Body mass index, mean (SD), kg/m<sup>2</sup> | 29.88 (6.01) | 29.20 (5.59) | 28.52 (5.31) | 27.90 (4.78) |
| Physical activity, mean (SD), MET-hours/w | 37.08 (39.46) | 42.98 (44.06) | 43.73 (39.08) | 47.96 (49.10) |
| Townsend deprivation index, No. (%) |  |  |  |  |
| Low | 102 (28.65) | 104 (29.21) | 140 (39.22) | 134 (37.64) |
| Middle | 118 (33.15) | 135 (37.92) | 115 (32.21) | 107 (30.06) |
| High | 136 (38.20) | 117 (32.87) | 102 (28.57) | 115 (32.30) |
| Smoking status, No. (%) |  |  |  |  |
| Never | 139 (39.04) | 167 (46.91) | 176 (49.30) | 186 (52.25) |
| Previous | 167 (46.91) | 146 (41.01) | 134 (37.54) | 124 (34.83) |
| Current | 45 (12.64) | 40 (11.24) | 44 (12.32) | 45 (12.64) |
| Alcohol consumption, No. (%) |  |  |  |  |
| Daily to three times a week | 119 (33.43) | 134 (37.64) | 109 (30.53) | 119 (33.43) |
| Twice a week to once a month | 114 (32.02) | 140 (39.33) | 142 (39.78) | 150 (42.13) |
| Never or special occasions only | 120 (33.71) | 81 (22.75) | 104 (29.13) | 87 (24.44) |

Abbreviations: MET, metabolic equivalent.

As shown in **Table 2**, higher IGF1 concentrations were associated with a reduced risk of COVID-19 mortality in the multivariable models (Model 1, OR comparing quartile 5 verse 1: 0.58, 95% CI: 0.39-0.85, *P*-trend=0.01). The association remained stable after further adjustment for the other confounders (Model 2, OR: 0.63, 95% CI: 0.43-0.93, *P*-trend=0.03).

**Table 2.**
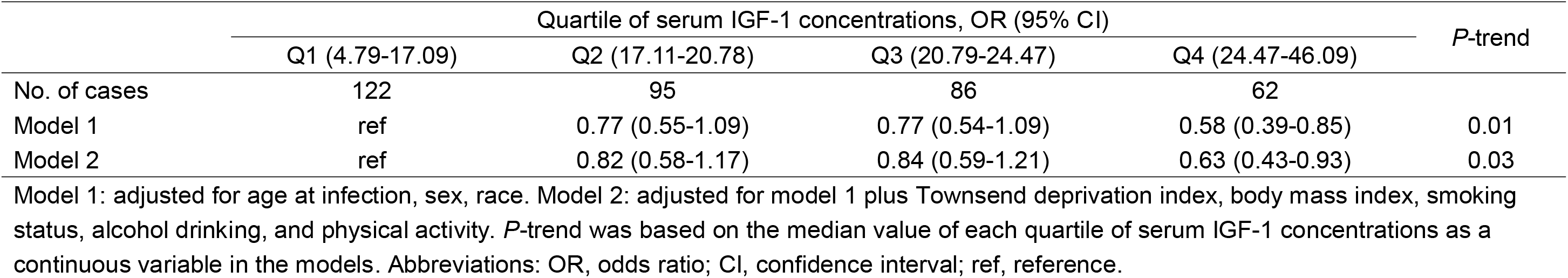
Association between serum IGF-1 concentrations and COVID-19 mortality.

**Figure 2** shows the forest plot results of stratified analyses. The inverse associations of IGF-1 with a COVID-19 mortality were largely consistent across subgroups, except for sex and smoking status. The association appeared to be stronger in women and nonsmokers (both *P*-interaction=0.01).

**Figure 2.**
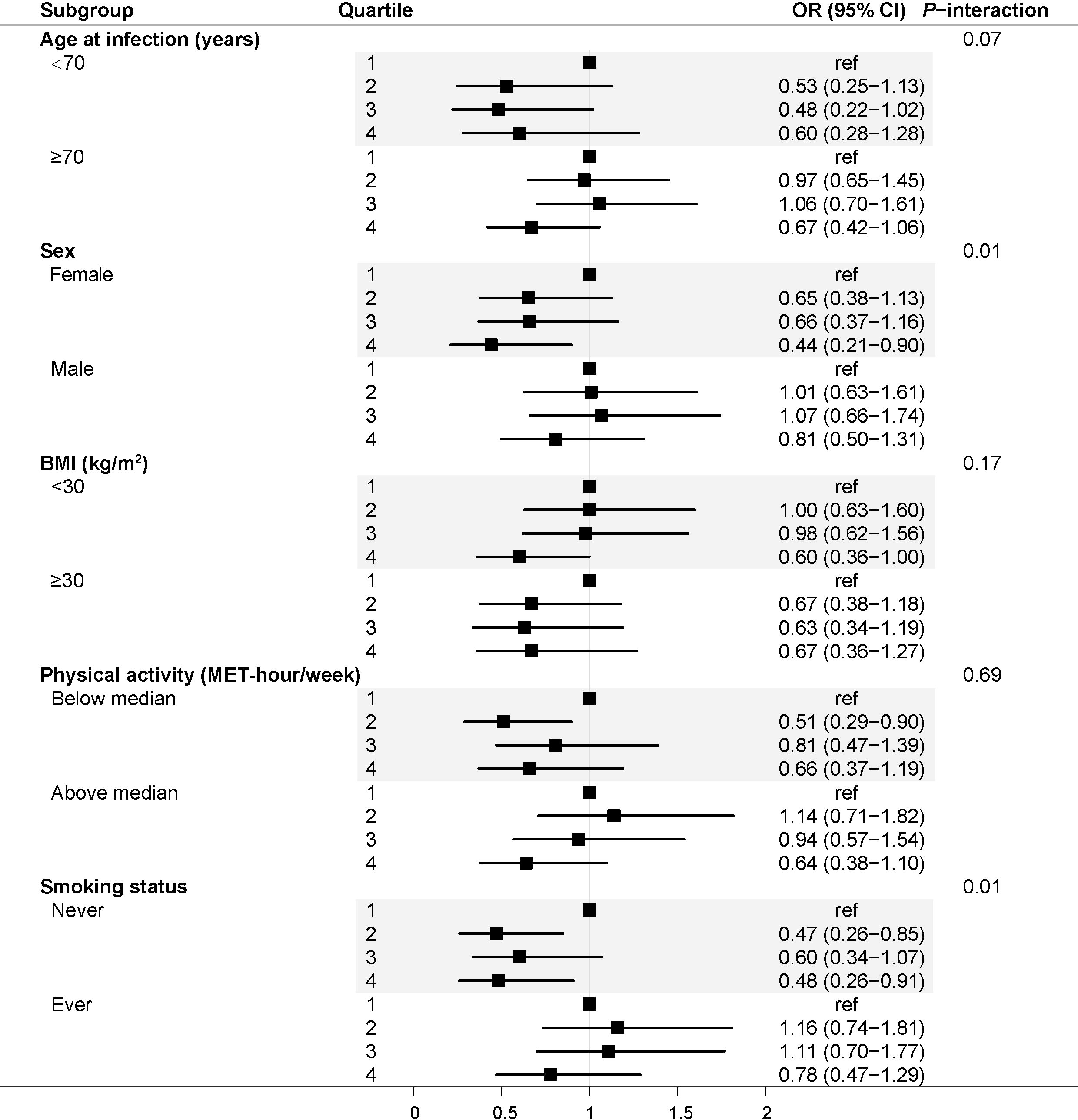
Forest plots of stratified analysis for the associations between serum IGF-1 concentrations and the risk of COVID-19 mortality.

Sensitivity analyses showed that the aforementioned associations remained basically unchanged after excluding 238 individuals who had baseline cancer or cardiovascular disease (**Table 3**).

**Table 3.**
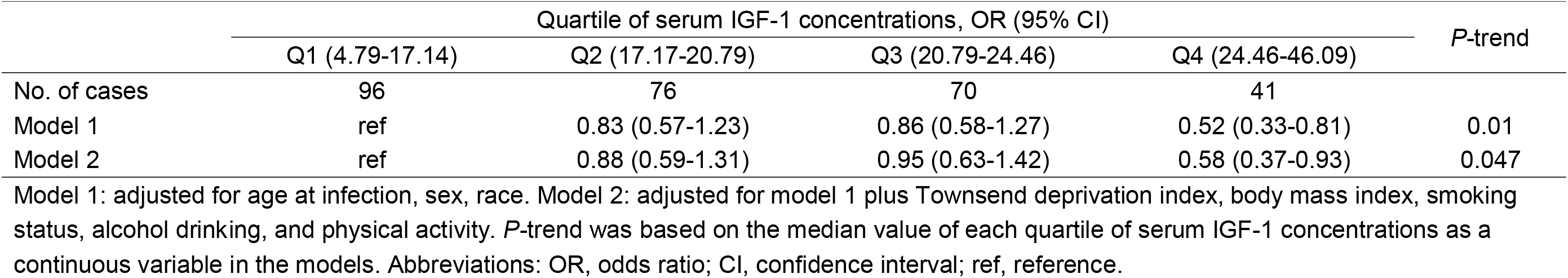
Association between serum IGF-1 concentrations and COVID-19 mortality after excluding participants with baseline cancer or vascular disease (n=238)

## Discussion

To the best of our knowledge, this study was the first to demonstrate an inverse association between pre-diagnostic circulating levels of IGF-1 and COVID-19 mortality risk among COVID-19 patients, particularly for women and nonsmokers. Our findings suggest that IGF-1 might play a role in COVID-19 prognosis.

Cytokine storm has been recognized as one of the major causes of ARDS and multiple organ failure in COVID-19 patients. Therefore, effectively suppressing the cytokine storm is important to prevent the disease deterioration and reduce COVID-19 mortality. Markedly elevated levels of circulating IL-6, IL-2R, IL-10, TNFα, etc. have been detected in patients with severe COVID-19 ^10 11^. These cytokines are mainly produced by macrophages and lymphocytes which have been implicated in cytokine storm ^12^. Several studies in humans and animal models have suggested an anti-inflammatory effect of IGF-1. For example, there is an inverse relationship between circulating IGF-1 and IL-6 levels ^13 14^, and IGF-1/IGF binding protein-3 administration to severely burned patients effectively attenuated inflammatory effects and reduced IL-6 levels ^15^. Recombinant human IGF-1 infusion into ApoE-deficient mice significantly decreased macrophage infiltration by downregulating IL-6 and TNF-α expression ^16^. In addition, recombinant human IGF-1 can mediate autoimmune suppression in mouse models of autoimmune disease by increasing regulatory T cells in affected tissues ^7^. However, it remains uncertain whether gender and smoking status modify the role of IGF-1 in immune regulation, and other studies also reported that IGF-1 might exert pro-inflammatory effects ^17 18^. Future studies are necessary to clarify the mechanism of IGF-1 in COVID-19 prognosis.

Our study has several strengths. First, UK Biobank is a well-designed large cohort and are providing reliable data on COVID-19 diagnosis and related deaths, allowing us to do the analysis in a timely fashion. Second, biochemistry assays of IGF-1 were performed in a single dedicated central laboratory by a standard, reliable method and strict quality control procedures. Third, we were able to adjust for various covariates on demographic and lifestyle factors.

Several potential limitations also need to be acknowledged. First, the observational nature of this study prevents us from inferring causality. However, our sensitivity analyses excluding baseline CVD and cancer supported the robustness of the findings. Second, given the lack of repeated IGF-1 measurements, we were unable to analyze the relationship between dynamic IGF-1 concentrations and COVID-19 mortality. However, we calculated the intraclass correlation coefficient (ICC) between IGF1 measurements collected 4 years apart in a subcohort (n= 16,356). The ICC value of 0.78, consistent with the previous data ^19^, indicates that IGF1 levels are generally stable over time. Third, due to limited coverage of coronavirus testing in the UK, ascertainment bias cannot be avoided. In addition, UK Biobank is not a representative sample of the UK population ^20^, limiting ability to generalize the results to the whole UK or other populations.

## Conclusions

The current study indicates that higher serum IGF-1 concentrations are associated with a lower risk of COVID-19 mortality. Further studies are warranted to validate our findings and clarify underlying mechanisms.

## Data Availability

The data that support the findings of this study are available from UK Biobank (https://www.ukbiobank.ac.uk/), but restrictions apply to their availability. These data were used under licence for the current study and so are not publicly available. The data are available from the authors upon reasonable request and with permission of UK Biobank.

http://biobank.ndph.ox.ac.uk/showcase/

## Acknowledgments

We are grateful to UK Biobank participants. This research has been conducted using the UK Biobank resource under application number 52217.

## Author Contributions

DH was responsible for the conception and design of the study. XF and DH had full access to all the data in the study and took responsibility for the integrity of the data and the accuracy of the data analysis. XF and CY did the statistical analysis and drafted the manuscript. JW, MY, HM, GJ, MS, ZH, and HS critically revised the manuscript for important intellectual content. All authors reviewed and approved the final manuscript.

## Abbreviations

COVID-19: coronavirus disease 2019
BMI: body mass index
MET: metabolic equivalent
ICD: international classification of diseases
OR: odds ratio
CI: confidence interval.

